# Analysis of Residual Risk and Recurrent Event Trends Following Acute Coronary Syndrome: A Cohort Study

**DOI:** 10.1101/2024.09.08.24313086

**Authors:** Timothy C. Shuey, Stephen J. Voyce, Laney K. Jones, Alicia Johns, Caroline deRichemond, Scott A. LeMaire, Braxton Lagerman, Shikhar Agarwal

**Affiliations:** Heart and Vascular Institute, Geisinger, Danville, PA; Heart and Vascular Institute, Geisinger, Scranton, PA; Department of Genomic Health, Research Institute, Geisinger, Danville, PA; Department of Population Health Sciences, Geisinger, Danville, PA; Department of Phenomics Analytics and Clinical Data Core, Geisinger, Danville, PA

## Abstract

**Background:** A comprehensive real-world analysis of residual risk factors for recurrent major adverse cardiovascular events (MACE) following hospital admission for acute coronary syndrome (ACS) is lacking. The objectives of this study were: 1) to describe population trends for outcomes, risk factors, and medication prescribing patterns post-ACS and 2) to identify factors associated with recurrent MACE.

**Methods:** A retrospective cohort study of 4,884 post-ACS patients admitted at a large integrated healthcare system between 2015-2021 was performed to investigate the relationship between recurrent MACE (ACS, cerebrovascular events, all-cause mortality, and unplanned revascularization), modifiable risk factor trends, and medical therapy prescribing patterns. Patients were separated into 2 cohorts based upon whether they experienced recurrent MACE following the initial hospitalization. Data were obtained via programmatic extraction from the electronic health record. Descriptive statistics were performed. Generalized linear models were used to assess risk factor trends and pairwise comparisons were performed between time points.

**Results:** Median length of follow-up after ACS was 31.2 months. Recurrent MACE occurred in 28% of patients. Despite 95.9% of all patients receiving prescriptions for high-intensity statins, >40% did not achieve LDL-C goal of <70 mg/dL, and only 11.6% and 2.6% of all patients were prescribed ezetimibe or proprotein convertase subtilisin kexin type 9 inhibiting monoclonal antibodies, respectively. Although >30.0% of patients had triglycerides ≥150 mg/dL at all time points, ≤6% were prescribed any non-statin triglyceride lowering therapy and 0.6% were prescribed icosapent ethyl. Persistent hypertriglyceridemia (≥150 mg/dL) was associated with recurrent MACE at 6-, 12-, and 24-months post-ACS (p<0.05), and the relative risk ranged between 1.20-1.35 at those timepoints.

**Conclusions:** This study demonstrates the need for more comprehensive post-ACS care to address residual cardiometabolic risk factors and suboptimal prescribing patterns for indicated therapies. Targeted strategies are needed to address hypertriglyceridemia for cardiovascular risk reduction.

**Clinical Perspective:** *What is new?:*

- This retrospective cohort study of post-acute coronary syndrome (ACS) patients addressed significant gaps in the literature by performing a comprehensive analysis of all major modifiable risk factors and medical therapy prescribing patterns to describe secular trends and identify factors associated with recurrent MACE.
- Although all modifiable risk factors were suboptimally controlled, only hypertriglyceridemia (>150 mg/dL) was significantly associated with recurrent MACE.
- Despite >95% of patients being on high-intensity statins, >40% of post-ACS patients did not achieve LDL-C goal of <70 mg/dL and there was suboptimal intensification of lipid-lowering therapies proven to reduce residual cardiovascular risk.

*What are the clinical implications?:*

- Targeted strategies are needed to address elevated LDL-C and hypertriglyceridemia in the post-ACS population.
- Implementation strategies to educate clinicians may help to improve medical therapy prescribing patterns for secondary prevention through treatment of cardiometabolic disease.

## Introduction

Coronary heart disease (CHD) is the leading cause of cardiovascular death and is associated with a high degree of morbidity and health care costs.^1–5^ ACS is the most feared sequela of CHD and is defined by acute myocardial ischemia which can manifest as ST-segment elevation myocardial infarction (STEMI), non-STEMI (NSTEMI), and unstable angina (UA).^1,6^ ACS is estimated to cause more than 1 million hospitalizations and 2.4 million deaths in the US annually.^6–8^ The global burden of modifiable cardiac risk factors continues to increase concurrent with morbidity and mortality due to cardiovascular disease.^1,2,4^

Guideline-directed medical therapy (GDMT) and lifestyle changes to affect modifiable cardiac risk factors are the cornerstones of post-ACS management to reduce residual risk for recurrent major adverse cardiovascular events (MACE).^1,2,4,9,10^ However, the literature has consistently shown that many post-ACS patients do not achieve adequate risk factor control and implementation of evidence-based guidelines is often suboptimal.^9–20^ Although many patients do not achieve guideline-directed lipid target thresholds, recent literature has also suggested that newer lipid lowering therapies, such as proprotein convertase subtilisin kexin type 9 inhibiting monoclonal antibodies (PCSK9-mAb) and icosapent ethyl (IPE), are underutilized. ^21^ Despite proven cardiovascular benefits, medications such as sodium-glucose cotransporter 2 inhibitors (SGLT2i) and glucagon-like peptide agonists (GLP-1RA), are underutilized in patients with type 2 diabetes and CHD.^2,22^

We sought to perform a comprehensive analysis of trends in modifiable cardiac risk factors, medical therapy prescribing patterns, and outcomes in a cohort of United States patients beginning at the time of hospitalization for ACS. The comprehensive nature of this study addresses significant gaps in the literature on post-ACS care. The objectives of this study were twofold: 1) to describe population trends for outcomes, risk factors, and medication prescribing patterns and 2) to identify factors associated with recurrent MACE.

## Methods

### Study Setting

This study was performed at Geisinger, a large integrated healthcare delivery system that includes 10 hospitals, 19 cardiology ambulatory clinics, 44 primary care clinics, and a health plan located in northeastern and central Pennsylvania, United States. Hospitals within the Geisinger healthcare system are Joint Commission/American Heart Association Comprehensive Heart Attack Centers and have received the American Heart Association’s Mission: Lifeline® STEMI Receiving Center Gold Plus recognition.

### Study Design

A retrospective cohort study was performed of patients who were admitted for ACS and had at least 6 months of subsequent outpatient follow-up data within the Geisinger health system. The patients were divided into 4 cohorts: 1) those who experienced recurrent 4-point MACE following the index hospitalization, 2) those with recurrent 3-point MACE, 3) those without recurrent 4-point MACE, and 4) those without recurrent 3-point MACE. Index hospitalization was defined as the first admission for ACS during the study period – this did not refer to the first lifetime event. We then compared the cohorts to identify risk factor trends and medication prescribing patterns that were associated with recurrent MACE.

### Outcomes

The primary outcome, 4-point MACE, was a composite of recurrent ACS, unplanned coronary revascularization, cerebrovascular events (ischemic stroke, hemorrhagic stroke, or transient ischemic attack), and all-cause mortality. Unplanned coronary revascularization was defined as a procedure >14 days after the index hospitalization and linked with an ACS diagnosis code. The secondary outcome, 3-point MACE, was a composite of ACS, cerebrovascular events, and all-cause mortality. For all analyses of modifiable risk factor trends and medication prescriptions, recurrent 4-point and 3-point MACE cohorts were compared to the cohorts without recurrent 4-point and 3-point MACE, respectively. Of note, there are shared patients between cohorts due to the overlapping definitions of MACE.

### Inclusion Criteria

Inclusion criteria for all patients were as follows: ≥18 years of age, treated for ACS within the Geisinger health system, and ≥1 primary care or outpatient cardiology follow-up encounter ≥6 months after the index admission. The index hospitalization and outpatient follow-up encounters needed to occur within the health system. The diagnostic criteria for STEMI, NSTEMI and UA used were per the Fourth Universal Definition of Myocardial Infarction. ^7^ STEMI and NSTEMI diagnosis codes needed to be associated with an inpatient or emergency department encounter. For STEMI and NSTEMI, coronary angiography ± revascularization must have occurred ≤14 days from the date of admission. This requirement was used to programmatically exclude encounters for type 2 myocardial infarction or nonischemic myocardial injury. The ICD-10 codes for type 2 myocardial infarction and nonischemic myocardial injury were first introduced in October 2017 and 2021, respectively, and it is known that these diagnoses have often been coded incorrectly as ACS.^23,24^ It was felt that patients with type 2 myocardial infarction or nonischemic myocardial injury would be less likely to have undergone invasive angiography. For patients diagnosed with UA (defined as having symptoms suggestive of ischemia with negative cardiac biomarkers), coronary revascularization was required ≤14 days from the date of admission. Overall, the criteria for coronary angiography ± revascularization was used to minimize ascertainment bias.

### Exclusion Criteria

Exclusion criteria were as follows: death during the index hospitalization, <6 months of outpatient follow-up data within the Geisinger health system, type 2 myocardial infarction, and nonischemic myocardial injury.

### Data Collection

All data were obtained exclusively from encounters between 1/1/2015-12/31/2021 via programmatic extraction from the electronic health record (EHR). One reviewer performed chart reviews to verify the accuracy of programmatic data collection. The index hospitalization for ACS had to occur between 1/1/2015-6/30/2021 to allow for a minimum of 6 months of outpatient follow-up data. Following ACS, we collected data points in 6-month intervals through 12/31/2021, which allowed for a potential maximum of 84 months of follow-up data. Length of follow-up was defined as the date of admission for the index inpatient encounter through the latest available data point of interest in the EHR. Lost to follow-up was defined as having no recorded data points in the EHR for ≥12 consecutive months.

Baseline past medical history data were extracted from 3 potential sources in the EHR: problem lists, diagnoses listed in ≥1 inpatient encounter, or diagnoses listed in ≥2 outpatient encounters. Past medical history and recurrent MACE were assessed via ICD 9 and 10 codes, CPT codes, and dates of decease from inpatient and outpatient encounters. All ICD diagnosis codes and CPT codes are listed in the supplementary materials.

We collected the following modifiable risk factor data: LDL-C, triglycerides (TG), HDL-C, non-HDL-C, hemoglobin A1c (HbA1c), body mass index (BMI), systolic blood pressure, and tobacco use status. Baseline data were collected during the index hospitalization (month 0). To account for the standard practice of measuring HbA1c at ≥3-month intervals, an exception was made which allowed HbA1c recorded ≤3 months before hospitalization to be included with baseline data. Following hospital discharge, modifiable risk factors were analyzed at 6-month intervals. If multiple data points for a risk factor were recorded ± 3 months from a 6-month interval, the value recorded closest to the 6-month interval was selected. For example, if systolic blood pressures were recorded at 10 months and 13 months post-ACS, the value recorded at 13 months would be included with the 12-month data and the value recorded at 10 months would be suppressed. Tobacco use status was assessed at baseline and the latest patient encounter.

Outpatient prescription data were analyzed at baseline followed by 6-month intervals for the following medication classes: beta-blockers, angiotensin-converting enzyme inhibitors (ACEi), angiotensin receptor blockers (ARB), statins, PCSK9-mAb, fibrates, IPE, oral antiplatelets, SGLT2i, GLP-1RA, metformin, and insulin.

### Statistical Methods and Data Analysis

Categorical outcomes were reported using frequencies and percentages. For continuous variables, medians and interquartile ranges were used for non-normal data. Descriptive statistics were presented for baseline demographics which were then compared by recurrent MACE status. The Wilcoxon rank sum test was used for continuous variables and Chi-square tests were performed for categorical variables. To examine the modifiable risk factors, generalized linear models with a lognormal distribution were utilized to assess the trend over 6-month intervals from baseline to 54 months. Pairwise comparisons with Tukey Kramer adjustments were used to examine differences between time points. Box and whisker plots were created to present the summary statistics at each time point. For LDL-C, the trend was also assessed for the subsets of patients who were statin-treated and statin-naïve; the percent at goal for these 2 subsets were also examined and compared at each time point. For TG and HbA1c, we also assessed trends for the subsets of nondiabetic patients, prediabetic patients and diabetic patients. For tobacco use status, the status was compared at baseline versus last known assessment using the McNemar test to assess overall differences in the statuses between the 2 time points. For each risk factor, we compared the percent at goal at each timepoint for the cohorts with and without recurrent MACE. The comparisons at each time point were tested using Chi square tests or Fisher’s exact tests. Additionally, relative risks were reported for TG. For LDL-C, we also examined the number of patients at goal at both baseline and 6 months. Similarly, the numbers of patients on each of the medications of interest were compared between the recurrent MACE cohort and those without recurrent events; Chi square tests and Fisher’s exact tests were also used for these comparisons. Kaplan Meier curves were utilized to assess both the survival probability and hazard rates for both recurrent MACE and all-cause mortality. Statistically significant differences were determined at the alpha level of 0.05. Analyses were conducted in SAS Enterprise Guide Version 8.2 (SAS Institute Inc., Cary NC, USA).

### Data Integrity and Regulatory Approval

The data that support the findings of this study are available from the corresponding author upon reasonable request. The first and corresponding author had full access to all data in the study and take responsibility for its integrity and data analysis. The institutional internal review board (IRB# 2021-0667) determined that this study was exempt. The STROBE guidelines were adhered to for this manuscript.^25^

## Results

### Study Population

The study population baseline characteristics and cohort construction are as shown in Table 1 and Figure 1. There were 4,884 patients overall. The number of patients by cohort were as follows: 1023 with recurrent 3-point MACE, 1382 with recurrent 4-point MACE, and 3502 without recurrent MACE. The median age was 64 years old (IQR 56, 73) and the population was predominately White (97.6%). Biological sex was 65.4% male and 34.6% female. Baseline past medical history was notable for: 22.7% with prior ACS, 5.6% with prior CVA, 55.1% with dyslipidemia, 23.2% with any form of diabetes mellitus, and 57.4% with hypertension. A comparison of baseline characteristics (Table 1) revealed significant differences between the cohorts with recurrent MACE and those without recurrent events. The cohorts with recurrent MACE were older (p<0.0001), had higher median HbA1c (p<0.0001), a greater incidence of cardiovascular comorbid conditions (p<0.0001), and a lower median left ventricular ejection fraction (LVEF) (3-point MACE: p<0.0001; 4-point MACE p=0.0008) with a greater percentage of patients with LVEF <35% (3-point MACE: p<0.0001; 4-point MACE p=0.0002). The cohorts with recurrent MACE had lower median LDL-C (p<0.0001) and a greater percentage of patients with LDL-C <70 mg/dL (p<0.0001), notwithstanding, there were no significant intercohort differences in the percentage of patients with statin prescriptions at baseline. Most notably, the cohorts with recurrent MACE had a greater percentage of patients with a prior history of ACS (p<0.0001).

**Figure 1.**
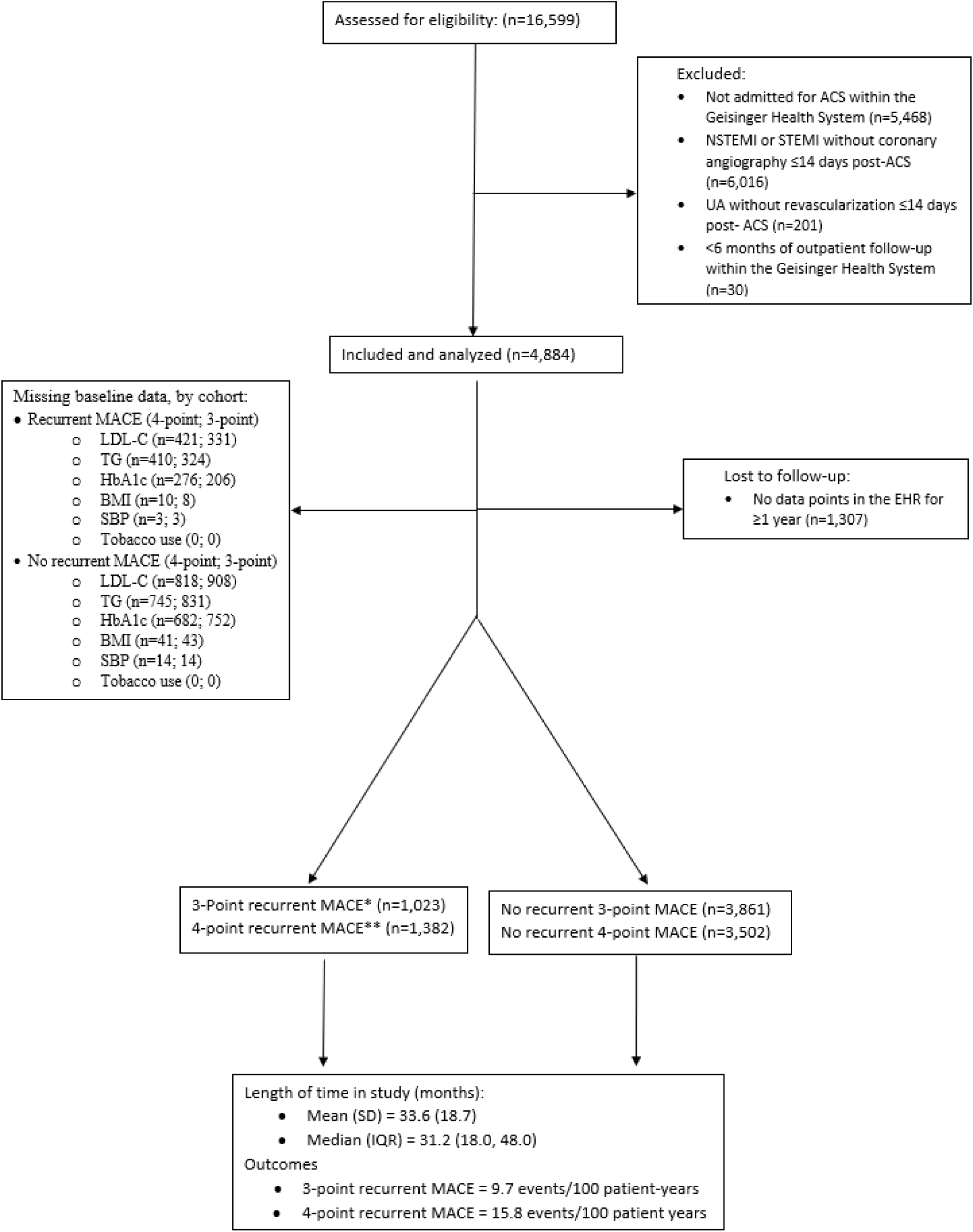
The STROBE diagram depicts the construction of the cohorts and key outcomes.

**Table 1.** Characteristics of Patients at Baseline.

|  | **Total Sample** | **Recurrent 3-point MACE** | **Recurrent 4-point MACE** | **No Recurrent 4-point MACE** |
| --- | --- | --- | --- | --- |
| Sample size (N) | (N = 4,884) | (N = 1,023) | (N = 1,382) | (N = 3,502) |
| Age in years, Mdn. (IQR) | 64 (56, 73) | 70 (60, 78)* | 68 (58, 76)* | 63 (55, 72) |
| Female sex — n (%) | 1,688 (34.6) | 377 (36.8) | 488 (35.1) | 1,200 (34.3) |
| Ethnicity — n (%)† |  |  |  |  |
| White | 4,769 (97.6) | 999 (97.7) | 1,350 (97.7) | 3,419 (97.6) |
| Body-mass index‡, Mdn. (IQR) | 29.8 (26.2, 34.3) | 29.6 (25.8, 34.4) | 29.8 (26.0, 34.3) | 29.8 (26.2, 34.2) |
| missing n = 51 |  |  |  |  |
| Systolic blood pressure, mm Hg, Mdn. (IQR) | 128 | 127.5* | 127.5* | 128 |
| missing n = 17 | (116.0, 140.0) | (115.0, 140.2) | (115.0, 140.0) | (116.0, 140.5) |
| At goal (<130 mmHg), n (%) | 2,627 (53.8) | 546 (53.5) | 746 (54.1) | 1,881 (53.9) |
| Lipid panel, Mdn. (IQR) |  |  |  |  |
| Non-HDL-C mg/dL | 124 | 116* | 119* | 125 |
| missing n = 1167 | (95.0, 158.0) | (87.0, 153.0) | (90.5, 156.0) | (97.0, 158.0) |
| HDL-C mg/dL | 40 | 39 | 39 | 40 |
| missing n = 1167 | (33.0, 49.0) | (33.0, 49.0) | (33.0, 49.0) | (33.0, 49.0) |
| At goal (>50, >40), n (%) | 1,443 (29.6) | 257 (37.0) | 356 (36.8) | 1087 (39.5) |
| LDL-C mg/dL | 96 | 88* | 91* | 98 |
| missing n = 1239 | (71.0, 126.0) | (63.0, 120.0) | (66.0, 123.0) | (73.0, 127.0) |
| At goal (<70 mg/dL), n (%) | 846 (17.3) | 214 (30.9)* | 270 (28.1)* | 576 (21.5) |
| Triglycerides mg/dL | 123 | 120 | 123 | 124 |
| missing n = 1155 | (85.0, 179.0) | (85.0, 178.0) | (87.5, 178.0) | (85.0, 180) |
| At goal (<150 mg/dL), n (%) | 2,387 (48.9) | 457 (65.4) | 628 (64.6) | 1,759 (63.8) |
| Hemoglobin A1c %, Mdn. (IQR) | 5.9 | 6.3* | 6.2* | 5.9 |
| missing n = 958 | (5.5, 7.1) | (5.6, 7.8) | (5.6, 7.7) | (5.5, 6.9) |
| At goal (<7.0%), n (%) | 2,842 (58.2) | 503 (61.6)* | 717 (64.8)* | 2,125 (75.4) |
| History of tobacco use — n (%) |  |  |  |  |
| missing n = 17 |  |  |  |  |
| Active or former tobacco use disorder | 3,189 (65.3) | 688 (67.2) | 937 (67.8) | 2,252 (64.3) |
| Never used tobacco | 1,678 (34.4) | 330 (32.3) | 439 (31.8) | 1,239 (35.4) |
| Cardiovascular history — n (%) |  |  |  |  |
| Diabetes mellitus | 1,135 (23.2) | 334 (32.6)* | 415 (30.0)* | 720 (20.6) |
| Dyslipidemia | 2,690 (55.1) | 711 (69.5)* | 902 (65.3)* | 1,788 (51.1) |
| Hypertension | 2,803 (57.4) | 756 (73.9)* | 941 (68.1)* | 1,862 (53.2) |
| HFrEF§ | 173 (3.5) | 90 (8.8)* | 97 (7.0)* | 76 (2.2) |
| HFpEF\|\| | 223 (4.6) | 108 (10.6)* | 115 (8.3)* | 108 (3.1) |
| Cerebrovascular events | 275 (5.6) | 130 (21.0)* | 145 (10.5)* | 130 (3.7) |
| Acute coronary syndrome | 1,108 (22.7) | 368 (36.0)* | 443 (32.0)* | 665 (19.0) |
| Ventricular tachycardia | 93 (1.9%) | 40 (3.9%)* | 44 (3.2%)* | 49 (1.4%) |
| LVEF# %, Mdn. (IQR) | 52 | 47* | 48* | 52 |
| missing n = 2,450 |  |  |  |  |
|  | (42.0, 57.0) | (37.0, 57.0) | (37.0, 57.0) | (42.0, 57.0) |
| LVEF<sup>#</sup> ≤35% — n (%) | 367 (7.5%) | 78 (22.0%)* | 107 (19.1%)* | 260 (13.9%) |

\*Indicates p<0.05. Recurrent 4-point and 3-point MACE cohorts were compared to the cohorts without recurrent 4-point and 3-point MACE, respectively.
†Ethnicity was self-reported data documented in electronic medical record
‡ The body-mass index is the weight in kilograms divided by the square of the height in meters
§ Heart failure with reduced ejection fraction
|| Heart failure with preserved ejection fraction

### MACE

Outcomes are presented in Table 2, Figure 1, and Figure 2. The survival and hazard curves for recurrent MACE are shown in Figure 4, and the survival curve for all-cause mortality is shown in Figure S5. The median length of follow-up was 31.2 months (IQR 18.0, 48). For the 4-point recurrent MACE cohort, the median time to the first repeat event was 14.0 months (IQR 3.5, 27.6); mean 18.0 months (SD 16.2). For the 3-point recurrent MACE cohort, the median time to the first repeat event was 17.8 months (IQR 7.8, 31.0); mean 21.1 months (SD 16.3). The greatest proportion of recurrent 3-point MACE was due to all-cause mortality (11.8%), while 4-point MACE was chiefly driven by unplanned coronary revascularization (13.7%). There were relatively equal proportions of MACE due to recurrent ACS (6.6%) and CVA (5.9%).

**Figure 2.**
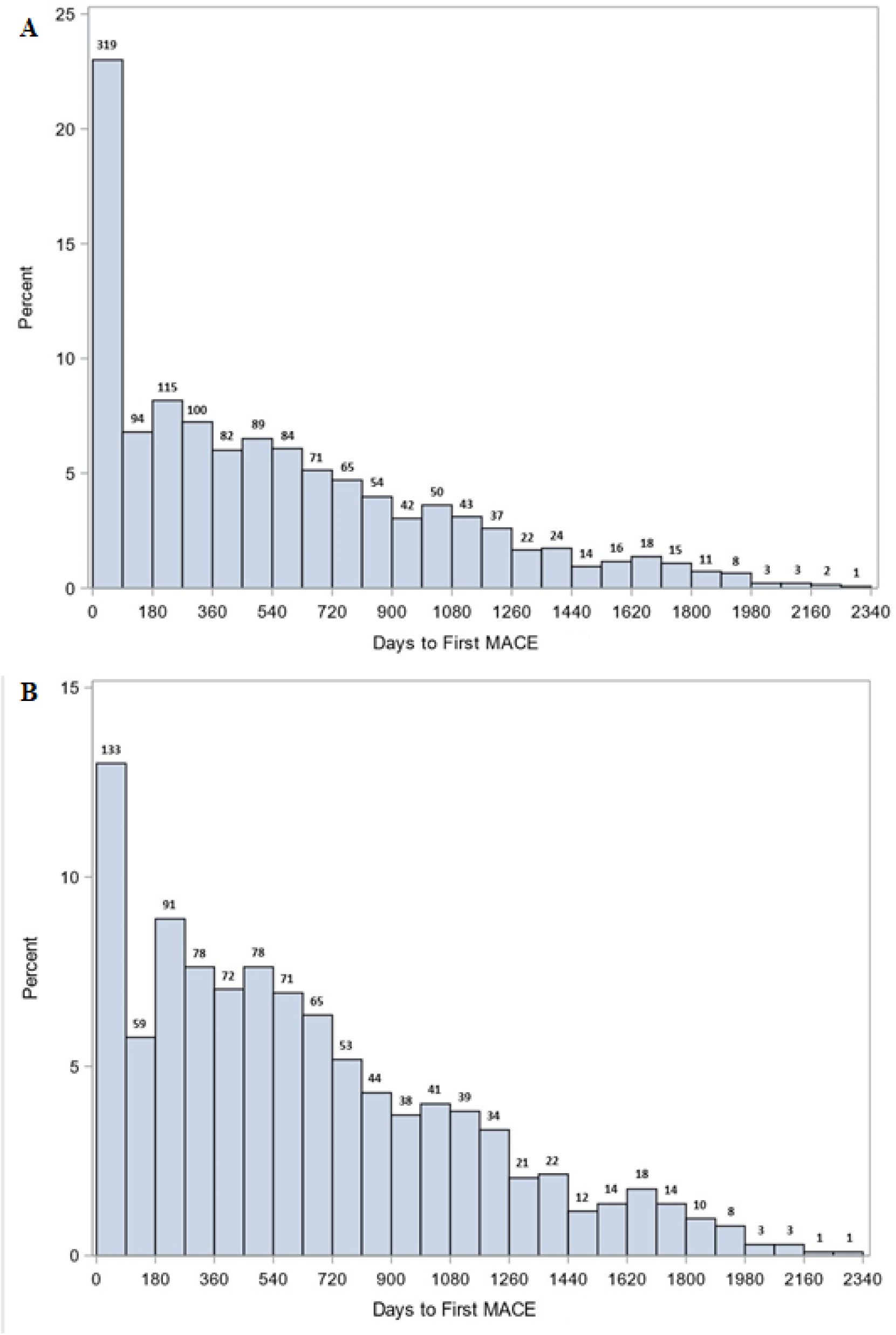
Days to first recurrent MACE histogram. Panel A is 4-point MACE. Panel B is 3-point MACE. Each bar represents 90-day timepoints. The number of events at each timepoint are shown above the respective bar.

**Figure 3.**
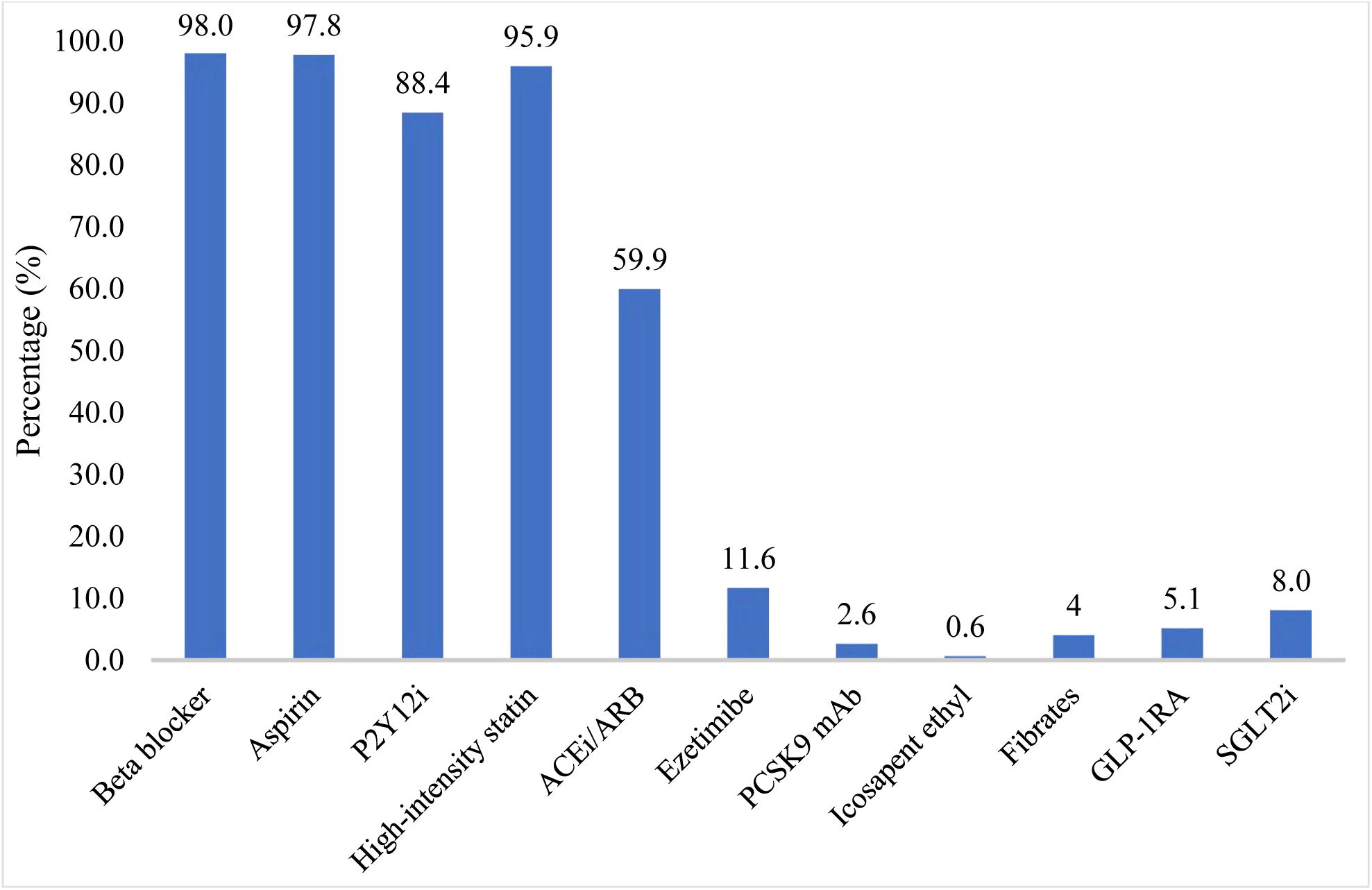
Prescription medications post-ACS.

**Figure 4.**
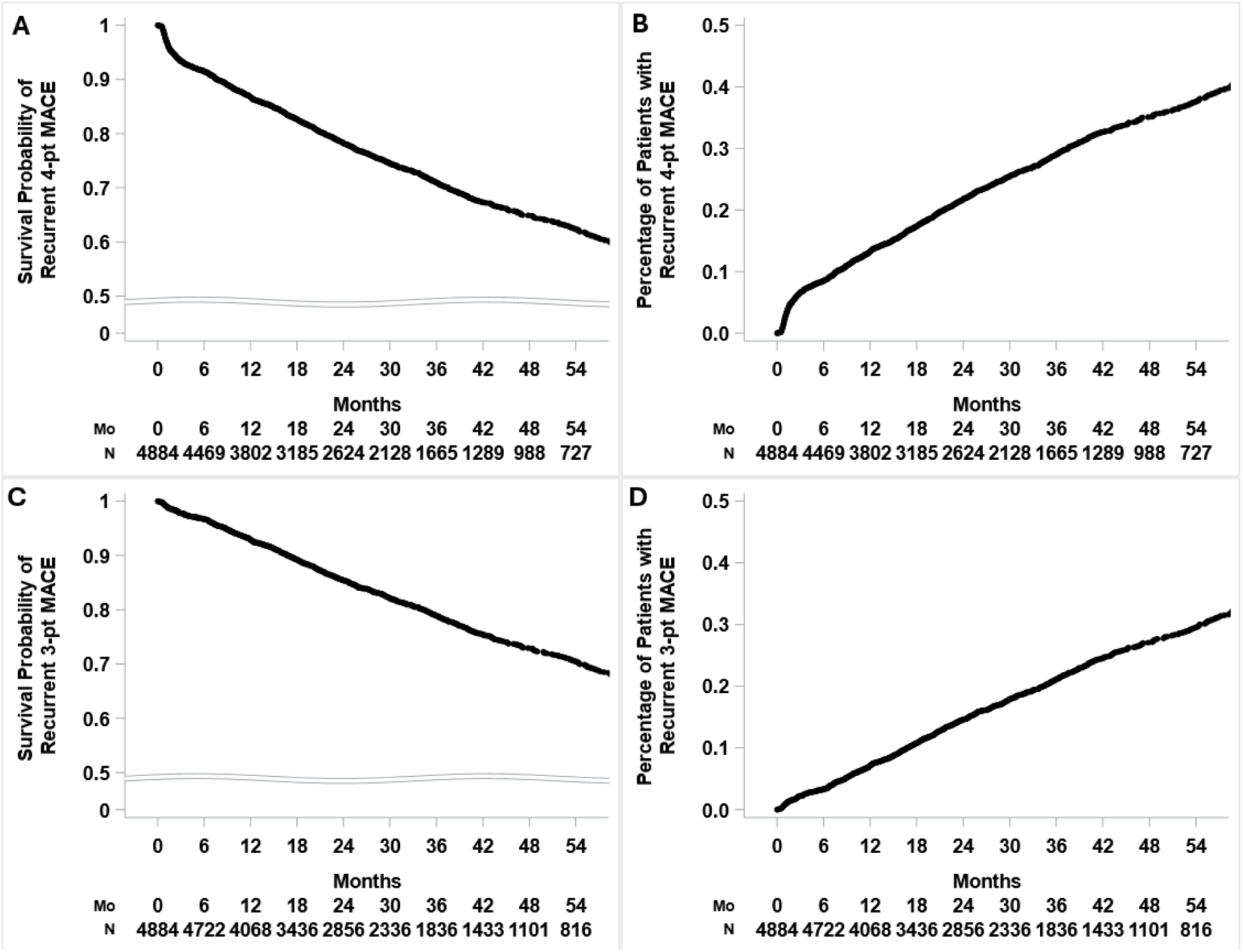
Survival and hazard curves for recurrent MACE. Panels A and B are the survival and hazard curves for 4-point MACE, respectively. Panels C and D are the survival and hazard curves for 3-point MACE, respectively. The tables below each curve show the number of patients at risk at each timepoint.

**Table 2.**
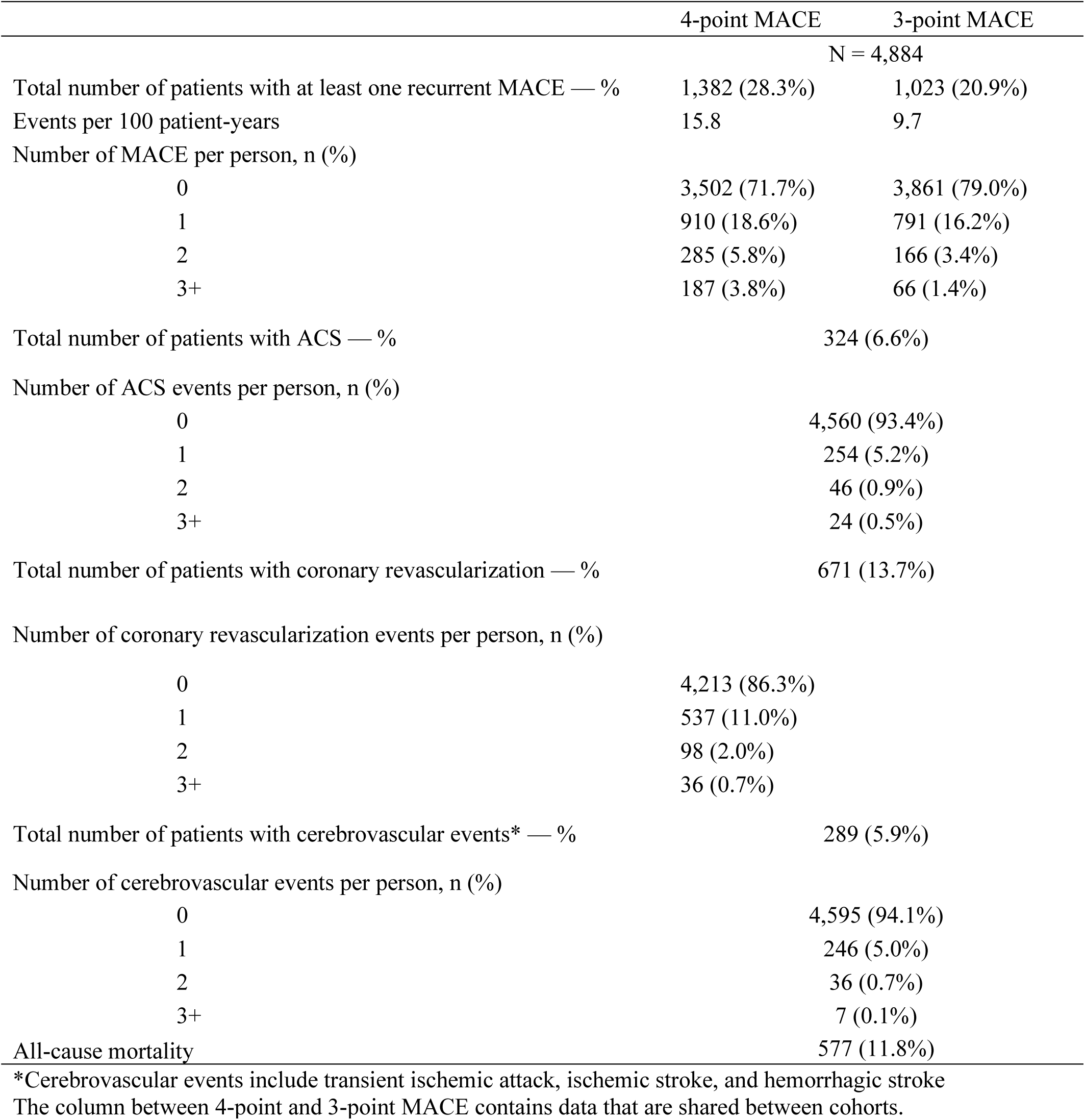
Incidence of major adverse cardiovascular events post-ACS.

### LDL-C

From baseline to 6 months post-ACS, the overall population had a median LDL-C decrease from 96 mg/dL (IQR 71, 126) to 66 mg/dL (IQR 50, 85). After 6-months, there was no significant change in median LDL-C (Figure S1) and >40% of patients had LDL-C above goal at all time points (Table 3). At baseline, 23.2% (846/3,645; missing = 1,239) of the overall population had LDL-C <70 mg/dL. At 6 months, 55.6% (990/1,779; missing = 3,105) of the overall population were at goal. At baseline, of those who ultimately experienced recurrent MACE, 32% (270/846) of patients had LDL-C <70 mg/dL compared to 24.7% (691/2,799) of patients who were not at goal. At 6 months post-ACS in those with recurrent MACE, 29% (292/990) had LDL-C <70 mg/dL compared to 30% (239/789) who were not at goal. Recurrent MACE was not a function of LDL-C ≥70 mg/dL (Table 3). To assess for a pooling effect, we performed a post-hoc analysis by comparing statin-naïve versus statin-treated patients, with the latter defined as having been on statin therapy at baseline (Figure S2). There was no statistically significant change in median LDL-C for either subset after 6 months post-ACS (Table S1).

**Table 3.** Cohort analysis: recurrent MACE as a function of LDL-C ≥70 mg/dL.

| Time period | 4-point MACE |  |  | 3-point MACE |  |  |
| --- | --- | --- | --- | --- | --- | --- |
| | $\geq 1$ Recurrent MACE | No Recurrent MACE | p-value | $\geq 1$ Recurrent MACE | No Recurrent MACE | p-value |
| Baseline | 71.9% (691/961) | 78.5% (2,108/2,684) | <0.0001 | 69.1% (478/692) | 78.6% (2,321/2,953) | <0.0001 |
| 6 months | 45.0% (239/531) | 44.1% (550/1,248) | 0.7153 | 45.2% (185/409) | 44.1% (604/1,370) | 0.6826 |
| 12 months | 46.4% (216/466) | 49.0% (501/1,023) | 0.3478 | 46.9% (152/324) | 48.5% (565/1,165) | 0.6137 |
| 18 months | 48.0% (196/408) | 48.8% (443/907) | 0.7875 | 49.7% (148/298) | 48.3% (491/1,017) | 0.6739 |
| 24 months | 44.6% (156/350) | 47.2% (359/760) | 0.4080 | 44.7% (106/237) | 46.8% (409/873) | 0.5609 |
| 30 months | 47.4% (127/268) | 49.1% (312/635) | 0.6316 | 49.5% (94/190) | 48.4% (345/713) | 0.7900 |
| 36 months | 46.0% (103/224) | 49.6% (265/534) | 0.3598 | 45.5% (71/156) | 49.3% (297/602) | 0.3946 |

Numbers in the table are percentage (n LDL-C $>70$ mg/dL / n LDL-C collected)

### Triglycerides

For the overall population, the median TG level at time of ACS was 123.0 mg/dL (IQR 85.0, 179.0) and there were no significant differences between cohorts. Throughout the study period, there was no statistically significant change in median TG level, and >30% of patients had TG above goal (>150 mg/dL) at all time points (Table 4). Persistent hypertriglyceridemia (TG >150 mg/dL) demonstrated a significant association with recurrent 4-point and 3-point MACE at 6, 12, and 24 months (p<0.05) and the relative risk ranged between 1.20-1.35 at those timepoints (Table 4 and Table S2).

**Table 4.** Cohort analysis: recurrent MACE as a function of TG ≥150 mg/dL.

| Time period | 4-point MACE |  |  | 3-point MACE |  |  |
| --- | --- | --- | --- | --- | --- | --- |
| | $\geq 1$ Recurrent MACE | No Recurrent MACE | p-value | $\geq 1$ Recurrent MACE | No Recurrent MACE | p-value |
| Baseline | 35.4% (344/972) | 36.2% (998/2,757) | 0.6519 | 34.6% (242/699) | 36.3% (1,100/3,030) | 0.4034 |
| 6 months | 37.1% (188/507) | 30.4% (366/1,205) | 0.0068 | 39.3% (152/387) | 30.3% (402/1,325) | 0.0009 |
| 12 months | 40.3% (179/444) | 32.1% (315/981) | 0.0026 | 41.7% (130/312) | 32.7% (364/1,113) | 0.0033 |
| 18 months | 37.2% (149/401) | 37.6% (335/892) | 0.8909 | 39.0% (114/292) | 37.0% (370/1,001) | 0.5185 |
| 24 months | 38.4% (132/344) | 32.2% (238/738) | 0.0480 | 40.1% (95/237) | 32.5% (275/845) | 0.0306 |
| 30 months | 40.8% (108/265) | 34.2% (214/626) | 0.0621 | 40.7% (77/189) | 34.9% (245/702) | 0.1379 |
| 36 months | 35.5% (77/217) | 34.3% (182/530) | 0.7655 | 34.2% (52/152) | 34.8% (207/595) | 0.8934 |

Numbers in the table are percentage (n TG $>150$ mg/dL / n TG collected)

### Hemoglobin A1c

Post-ACS trends in HbA1c were analyzed for the following groups: total population, nondiabetic (HbA1c <5.7%), prediabetic (HbA1c 5.7-6.4%), and T2DM (HbA1c ≥6.5%) (Table S3). The HbA1c levels that defined each group are per the American Diabetes Association diagnostic criteria. Patients with HbA1c in the nondiabetic and prediabetic ranges at baseline that progressed to new onset T2DM are shown in Figure S3. An analysis of unique patients found that 4.3% (59/1,364) of nondiabetic and 23.1% (267/1,158) of prediabetic patients eventually developed T2DM over a median follow-up of 31.2 months. The recurrent MACE cohorts had a significantly greater proportion of patients with HbA1c >7.0% (p<0.0001) at baseline determination (Table 1). At all subsequent time points, the recurrent MACE cohorts had a greater proportion of patients with HbA1c >7.0%, but this trend did not reach statistical significance.

### Body Mass Index

BMI was static post-ACS with the median approximating 30.0 kg/m^2^. The recurrent MACE cohorts had a statistically lower proportion of patients with BMI >30 kg/m^2^ at multiple time points (Table S4).

### Systolic Blood Pressure

The median systolic blood pressure ranged between 126.0-128.0 mm Hg (Figure S4). When the percentage of patients in each cohort with a systolic blood pressure >130 mm Hg were compared, there were no significant differences at any timepoint. Systolic blood pressure was >130 mm Hg for >35% of patients in all cohorts at all time points.

### Tobacco Use Status

Over 65% of patients in all cohorts were either active tobacco users or had a history of former use. For all cohorts, there was an approximate 10% decrease in active tobacco users between baseline determination and last known assessment.

### Medication Prescription Trends

For the overall population, the percentage of patients with post-ACS prescriptions by medication class are presented in Figure 3. Medical therapy prescribing patterns were analyzed as follows: 1) the overall population to describe secular trends in post-ACS care and 2) by cohort to assess if those with recurrent MACE received similar medical therapy in comparison to those without recurrent events. Standard of care for post-ACS patients includes a high-intensity statin, beta-blocker, aspirin, and a P2Y12 inhibitor: an analysis of unique patients showed that 34.3% (1,677/4,884) were on this combination; the cohorts with recurrent MACE had a significantly greater proportion of patients on this combination at 6 months post-ACS and at all subsequent time points (p<0.0001). The AHA/ACC multisociety guideline on the management of blood cholesterol recommends the addition of ezetimibe to maximally tolerated statin therapy when the LDL-C level remains ≥70 mg/dL in the setting of very-high risk atherosclerotic cardiovascular disease (ASCVD).^14^ An analysis of unique patients showed that 2.4% (119/4884) were on a combination of ezetimibe, high-intensity statin, beta-blocker, aspirin, and a P2Y12 inhibitor; the cohort with recurrent MACE had a significantly greater proportion of patients (p<0.05) on this combination at multiple timepoints. However, there were <35 patients per cohort with an active prescription for this combination (ezetimibe, high-intensity statin, beta-blocker, aspirin, and a P2Y12 inhibitor) at each timepoint. Of note, an analysis of unique patients found that 3.0% (146/4884) of all patients were prescribed the combination of a statin and fibrate; when cohorts were compared by recurrent MACE status, there were no significant differences in the proportion of patients on this combination (p>0.05). For all cohorts, the proportion of patients on each individual medication class increased between 0 and 6 months post-ACS. An analysis of individual medication classes by cohort showed that those with recurrent MACE had a significantly greater proportion of patients (p<0.05) on aspirin, beta-blockers, P2Y12 inhibitors, and insulin. There were no significant differences between cohorts for the proportion of patients on the following individual medication classes: high-intensity statin, PCSK9-mAb, ezetimibe, SGLT2i, GLP1-RA, IPE, fibrates, and metformin. For fibrates, 4.0% (195/4884) of all patients were prescribed this medication class, with or without concomitant statin therapy, during the post ACS period. For IPE, only 0.6% (31/4,884) of all patients were prescribed this medication despite guideline recommendations and FDA indication in a high-risk population on statin therapy. High-intensity statins were prescribed to 95.9% of patients. SGLT2i are indicated for patients with a LVEF <50%; although ≥35% of all patients had a LVEF <50% at all time points, less than 5% of these patients were prescribed an SGLT2i at all time points. Patients with T2DM and CHD have an indication for early initiation of GLP-1RA to reduce the risk of MACE, yet <10% of eligible patients were prescribed this medication class at any time point.

## Discussion

A comprehensive analysis of post-ACS patients was performed to describe secular trends in risk factors and medical therapy prescribing patterns and to identify factors associated with residual risk for recurrent MACE. The key findings include undertreated LDL-C, an association between hypertriglyceridemia and recurrent MACE, the incidence of new onset T2DM post-ACS, and under-prescribing of indicated medical therapies (ezetimibe, PCSK9-mAb, IPE, GLP-1RA, and SGLT2i). Of note, 3.0% (146/4884) of patients were prescribed a combination of fibrate and statin therapy despite evidence-based guidelines categorically not supporting this strategy for risk reduction.^14^ Although hypertriglyceridemia was the only risk factor that demonstrated a significant association with recurrent MACE in these data, undertreated LDL-C and T2DM are well-established risk factors. The lack of association between recurrent MACE and LDL-C and T2DM are likely attributable to a median follow-up limited to 31.2 months.

Although prior investigations related to secondary prevention in the post-ACS population have described trends in cardiac risk factors, medical therapy, and outcomes, these investigations were limited in various ways, including: clinical trial study populations, pooling of multiple forms of ASCVD, limited in scope by studying select risk factors or medications, utilization of mathematical modeling in lieu of real-world data, reliance on clinician and patient survey data, short study durations, and studying non-US patient populations. ^9–13,15–21,23,26–37^ We addressed this gap in the literature by performing a comprehensive analysis of post-ACS patients to describe secular trends in risk factors and medical therapy prescribing patterns and to identify factors associated with residual risk for recurrent MACE. The key findings include undertreated LDL-C, an association between hypertriglyceridemia and recurrent MACE, the incidence of new onset T2DM post-ACS, and under-prescribing of indicated medical therapies (ezetimibe, PCSK9-mAb, IPE, GLP-1RA, and SGLT2i).

### Outcomes

There was a high incidence of recurrent events with 28.3% (1382/4,884) and 20.9% (1023/4,884) of patients experiencing 4-point and 3-point MACE, respectively, over a median follow-up of 31.2 months (Table 2). Nearly 10% of patients with recurrent events had >1 MACE during follow-up (Table 2). Prior real-world investigations into recurrent MACE in the post-ACS population have reported a wide incidence range, 11-48.4%, which is likely a function of variable definitions of MACE and different follow-up durations. ^18–20,34^

### Modifiable risk factors

Between 0 and 6 months post-ACS, the median LDL-C decreased from 96 mg/dL to 66 mg/dL for the total population. However, after 6 months there was no significant change in the population median LDL-C, and >40% of patients had LDL-C >70 mg/dL at all time points. The lack of a significant change in median LDL-C after 6 months suggests that medications were not titrated beyond initial in-hospital management. These findings align with prior investigations that demonstrated suboptimal management of cholesterol.^9,10,12–14,16–19,26–31,33,36^ The lack of statistically significant differences in LDL-C trends between cohorts suggests that other residual risk factors may have influenced the recurrent MACE rates observed in this study.

For patients with CHD on statin therapy with LDL-C at goal, elevated triglyceride levels are an independent marker for residual risk for ischemic events, which has been demonstrated in epidemiologic and mendelian randomization studies.^38–43^ In this investigation, hypertriglyceridemia (TG >150 mg/dL) demonstrated a significant association with recurrent 4-point and 3-point MACE. As shown in Table 4 and Table S2, the relative risk ranged from 1.20-1.35 at the timepoints that reached statistical significance (p<0.05). Despite trial data demonstrating a reduced risk of recurrent MACE in patients with hypertriglyceridemia post-ACS, only 0.6% of patients in this study were prescribed IPE.^43^ At the time of the initial encounter for ACS, we identified 1,342 unique patients with TG >150 mg/dL which persisted throughout the study period. This group represents a significant percentage of patients who may benefit from therapy with IPE.

T2DM is a key modifiable risk factor, often coupled with high BMI and sedentary lifestyle, and it has been shown that even elevated fasting plasma glucose in the prediabetic range portends elevated CVD risk. ^1,2,9,11,22,37,39,44–46^ There was a statistically significant progression to T2DM for patients who were nondiabetic and prediabetic patients at baseline (p<0.0001), and this represents an important care improvement opportunity for this population.^4,16–18,44–46^

BMI was persistently elevated at around 30 kg/m^2^ for the overall population and did not significantly change throughout the study period. Obesity is an often-uncontrolled risk factor in the post-ACS population.^10,16,17,20^ The cohorts with recurrent MACE had a significantly greater proportion of patients with BMI <30 kg/m^2^. This could potentially be explained by comorbid conditions associated with low BMI which could include, end-stage COPD, advanced heart failure, malignancy, and many other chronic conditions.

Systolic blood pressure was above goal for >33% of patients and did not significantly vary in either cohort throughout the study. These data suggest suboptimal intensification of antihypertensive therapy for many patients.

Over 65% of the entire population were active or former tobacco users at baseline. The proportion of patients who quit using tobacco during the study period was approximately 10% for all cohorts. Tobacco cessation remains a significant care gap for the post-ACS population.

### Medical therapy

The efficacy of medical therapies analyzed in this study (Figure 3) is established by a multitude of randomized clinical trials and are recommended by professional society guidelines.^8,11,14,21,22,35,43,44^ However, there are limited real-world data on newer medications for management of comorbid disease and secondary prevention for ACS, such as PCSK9-mAb, IPE, GLP1-RA, and SGLT2i.^21,22,26,27,33,45,46^

Medication prescribing patterns showed that patients with recurrent MACE did not uniformly receive a lower standard of care, in terms of medication prescriptions, compared to those without recurrent MACE. The cohort with recurrent MACE had a greater proportion of patients on guideline-directed medical therapy. The difference in outcomes between cohorts was not driven by medical prescribing patterns. However, for the total population, only 34.3% (1677/4884) were on standard of care (P2Y12i, aspirin, beta blocker, and high-intensity statin) and modifiable risk factors were not well-controlled. Despite 95.9% utilization of high-intensity statins, >40% of post-ACS patients did not achieve LDL-C goal of <70 mg/dL, but only 11.6% and 2.6% of all patients were on ezetimibe or PCSK9-mAb, respectively. Although 34.1% of patients had TG >150 mg/dL at 36 months post-ACS, <6% of the overall population were on non-statin TG lowering therapy and, specifically, only 0.6% were on IPE which has been shown to decrease risk for recurrent events in this population.^43^ Fibrate therapy is not supported by evidence-based guidelines, nor is this medication class indicated by the FDA, for cardiovascular risk reduction as monotherapy or as adjunctive lipid lowering therapy with statins.^14^ Despite this, 3.0% (146/4884) were prescribed a combination of statin and fibrate and 4.0% (195/4,884) were on a fibrate (with or without concomitant lipid lowering therapy)._ These data align with prior studies which found suboptimal intensification of lipid lowering therapy and underutilization of other indicated medications in similar very high-risk populations as defined by the AHA/ACC multisociety guideline on cholesterol management. ^10,13,14,16,17,20,21,26,28,30,33^

Since 2016, it has been known that GLP-1RA reduce the risk of recurrent MACE and all-cause mortality in patients with ASCVD and T2DM, and this has been reflected in European guidelines since 2019.^47–49^ There are recent trial data on semaglutide which showed a mortality benefit and decrease in recurrent MACE for patients with ASCVD and BMI ≥27 kg/m2 without T2DM, which lead to FDA approval for this indication on 3/8/2024.^50^ GLP-1RA have been indicated for weight management in patients with BMI ≥30 kg/m2 since liraglutide was approved for this indication in 2014. Despite this study population having a median BMI of 30 kg/m^2^ and 31.9% with T2DM, only 5.1% received a prescription for a GLP-1RA. Of note, most of the trial data and guidelines pertaining to GLP-1RA use in patients with CHD has been contemporaneous with the current study. This underscores the underutilization of GLP-1RA in the post-ACS population with comorbid cardiometabolic disease.

### Limitations

There are limitations inherent to a nonrandomized retrospective study design that preclude the ability to prove causation. As described above, undetermined confounding variables (e.g., comorbidities such as advanced malignancy or end-stage COPD) are a potential explanation for BMI <30 kg/m^2^ showing a statistically significant association with increased mortality. A subset analysis of BMI to assess for confounding comorbidities was not performed. Reliance on billing codes and problem list diagnoses is another potential source of error.^24^

As described in the methods section, the inclusion criteria required patients with UA to have undergone revascularization ≤14 days from the date of admission to facilitate excluding patients admitted with a provisional diagnosis of UA who underwent angiography but were found to have no or minimal coronary artery disease. This criterion is a limitation due to the potential for excluding patients with disease not amenable to revascularization or those patients who declined revascularization in favor of medical management. This criterion was used because it was felt that patients admitted with suspected UA but who were ultimately found to have no or minimal coronary artery disease, may have other noncardiac etiologies for chest pain and would likely outnumber those with UA and unrevascularized disease who were medically managed.

Tobacco use status is often documented using free text in notes instead of updating the categorical data in social history section of the EHR. Free text is not amenable to programmatic extraction for data mining without natural language processing. Therefore, we extracted tobacco use data from the social history section of the EHR at the index hospitalization and the latest available clinical encounter.

Data availability was limited to patients who routinely follow-up within this Geisinger health system. Medication data is particularly susceptible to inaccuracies given that patients may fill prescriptions at outside pharmacies and follow with clinicians outside this health system. Medication adherence was not assessed. There were a total of 1,307 patients meeting inclusion criteria who were lost to follow-up.

The COVID-19 pandemic may have altered outcomes data for morbidity and mortality.

## Conclusions

This investigation identified multiple targets for further investigation including an association between hypertriglyceridemia and MACE. Medical therapy optimization to address cardiometabolic residual risk remains an ongoing care gap for many post-ACS patients.

## Data Availability

The data that support the findings of this study are available from the corresponding author upon reasonable request. The first and corresponding author had full access to all data in the study and take responsibility for its integrity and data analysis.

## Acknowledgments

We would like to thank the staff at the Geisinger Health Sciences Library for assisting with the initial literature search.

## Disclosures

Timothy C. Shuey has no disclosures.

Alicia Johns has no disclosures.

Braxton Lagerman has no disclosures.

Laney K. Jones is a consultant for Novartis.

Caroline deRichemond has no disclosures.

Scott LeMaire serves as a consultant for Cerus and has served as a principal investigator for clinical studies sponsored by Terumo Aortic and CytoSorbents.

Shikhar Agarwal serves as a proctor for Edwards Lifesciences and as a speaker for Abbott Laboratories and Medtronic, Inc.

Stephen J. Voyce receives study funding from Astra Zeneca and Amarin Pharma, Inc.

## Supplementary Materials

Tables S1-S4

Figures S1-S5

## References

1. Tsao CW, Aday AW, Almarzooq ZI, et al. Heart Disease and Stroke Statistics-2022 Update: A Report From the American Heart Association. Circulation. 2022;145(8):e153–e639. doi:10.1161/CIR.0000000000001052

2. Vaduganathan M, Mensah GA, Turco JV, Fuster V, Roth GA. The Global Burden of Cardiovascular Diseases and Risk: A Compass for Future Health. J Am Coll Cardiol. 2022;80(25):2361–2371.

3. Mensah GA, Roth GA, Fuster V. The Global Burden of Cardiovascular Diseases and Risk Factors: 2020 and Beyond. J Am Coll Cardiol. 2019;74(20):2529–2532.

4. Roth GA, Mensah GA, Johnson CO, et al. Global Burden of Cardiovascular Diseases and Risk Factors, 1990-2019: Update From the GBD 2019 Study. J Am Coll Cardiol. 2020;76(25):2982–3021.

5. Summary of Global Burden of Disease Study Methods. J Am Coll Cardiol. 2022;80(25):2372–2425.

6. Reed GW, Rossi JE, Cannon CP. Acute myocardial infarction. Lancet. 2017;389(10065):197– 210.

7. Thygesen K, Alpert JS, Jaffe AS, et al. Fourth Universal Definition of Myocardial Infarction (2018). Circulation. 2018;138(20):e618–e651.

8. Bhatt DL, Lopes RD, Harrington RA. Diagnosis and Treatment of Acute Coronary Syndromes: A Review. JAMA. 2022;327(7):662–675. doi:10.1001/jama.2022.0358

9. Farkouh, Michael E., MD, MSc, Boden WE, MD, Bittner, Vera, MD, MSPH, et al. Risk Factor Control for Coronary Artery Disease Secondary Prevention in Large Randomized Trials. Journal of the American College of Cardiology. 2013;61(15):1607–1615. doi:10.1016/j.jacc.2013.01.044

10. Kotseva K, de Backer G, De Bacquer D, et al. Lifestyle and impact on cardiovascular risk factor control in coronary patients across 27 countries: Results from the European Society of Cardiology ESC-EORP EUROASPIRE V registry. European journal of preventive cardiology. 2019;26(8):824–835. doi:10.1177/2047487318825350

11. Farkouh ME, Domanski M, Sleeper LA, et al. Strategies for Multivessel Revascularization in Patients with Diabetes. The New England journal of medicine. 2012;367(25):2375–2384. doi:10.1056/NEJMoa1211585

12. Silva V, Matos Vilela E, Campos L, et al. Suboptimal control of cardiovascular risk factors in myocardial infarction survivors in a cardiac rehabilitation program. Revista portuguesa de cardiologia. 2021;40(12):911–920. doi:10.1016/j.repc.2021.01.015

13. Gitt AK, xel H, Feely J, et al. Persistent lipid abnormalities in statin-treated patients and predictors of LDL-cholesterol goal achievement in clinical practice in Europe and Canada. European journal of preventive cardiology. 2012;19(2):221–230. doi:10.1177/1741826711400545

14. Grundy SM, Stone NJ, Bailey AL, et al. 2018 AHA/ACC/AACVPR/AAPA/ABC/ACPM/ADA/AGS/APhA/ASPC/NLA/PCNA Guideline on the Management of Blood Cholesterol: A Report of the American College of Cardiology/American Heart Association Task Force on Clinical Practice Guidelines. Circulation. 2019;139(25):e1082–e1143.

15. Mehta RH, Bhatt DL, Steg PG, et al. Modifiable risk factors control and its relationship with 1 year outcomes after coronary artery bypass surgery: insights from the REACH registry. European Heart Journal. 2008;29(24):3052–3060. doi:10.1093/eurheartj/ehn478

16. Vedin O, Hagström E, Stewart R, et al. Secondary prevention and risk factor target achievement in a global, high-risk population with established coronary heart disease: baseline results from the STABILITY study. European journal of preventive cardiology. 2013;20(4):678– 685. doi:10.1177/2047487312444995

17. Bhatt DL, Steg PG, Ohman EM, et al. International Prevalence, Recognition, and Treatment of Cardiovascular Risk Factors in Outpatients With Atherothrombosis. JAMA: the journal of the American Medical Association. 2006;295(2):180–189. doi:10.1001/jama.295.2.180

18. Okkonen M, Havulinna AS, Ukkola O, et al. Risk factors for major adverse cardiovascular events after the first acute coronary syndrome. Annals of Medicine. 2021;53(1):817. doi:10.1080/07853890.2021.1924395

19. Simard T, Jung RG, Di Santo P, et al. Modifiable Risk Factors and Residual Risk Following Coronary Revascularization: Insights From a Regionalized Dedicated Follow-Up Clinic. *Mayo Clinic Proceedings.Innovations*, Quality & Outcomes. 2021;5(6):1138–1152.

20. Ferrari R, Ford I, Greenlaw N, et al. Geographical variations in the prevalence and management of cardiovascular risk factors in outpatients with CAD: Data from the contemporary CLARIFY registry. European journal of preventive cardiology. 2015;22(8):1056–1065. doi:10.1177/2047487314547652

21. Shen M, Aghajani Nargesi A, Nasir K, Bhatt DL, Khera R. Contemporary National Patterns of Eligibility and Use of Novel Lipid-Lowering Therapies in the United States. JAHA. 2022;11(18). doi:10.1161/jaha.122.026075

22. McCoy RG, Van Houten HK, Karaca-Mandic P, Ross JS, Montori VM, Shah ND. Second-Line Therapy for Type 2 Diabetes Management: The Treatment/Benefit Paradox of Cardiovascular and Kidney Comorbidities. Diabetes Care. 2021.

23. Martin LM, Januzzi JLJ, Thompson RW, et al. Clinical Profile of Acute Myocardial Infarction Patients Included in the Hospital Readmissions Reduction Program. Journal of the American Heart Association. 2018;7(16):e009339.

24. Diaz-Garzon J, Sandoval Y, Smith SW, et al. Discordance between ICD-Coded Myocardial Infarction and Diagnosis according to the Universal Definition of Myocardial Infarction. Clin Chem. 2017;63(1):415–419.

25. von Elm E, Altman DG, Egger M, et al. The Strengthening the Reporting of Observational Studies in Epidemiology (STROBE) statement: guidelines for reporting observational studies. Lancet. 2007;370(9596):1453–1457.

26. Mackinnon ES, Har B, Champsi S, et al. Guideline LDL-C Threshold Achievement in Acute Myocardial Infarction Patients: A Real-World Evidence Study Demonstrating the Impact of Treatment Intensification with PCSK9i. Cardiology & Therapy. 2023.

27. Cannon CP, de Lemos JA, Rosenson RS, et al. Getting to an ImprOved Understanding of Low-Density Lipoprotein-Cholesterol and Dyslipidemia Management (GOULD): Methods and baseline data of a registry of high cardiovascular risk patients in the United States. Am Heart J. 2020;219:70–77.

28. Zheutlin AR, Derington CG, Herrick JS, et al. Lipid-Lowering Therapy Use and Intensification Among United States Veterans Following Myocardial Infarction or Coronary Revascularization Between 2015 and 2019. Circulation.Cardiovascular Quality & Outcomes. 2022;15(12):e008861.

29. Colantonio LD, Huang L, Monda KL, et al. Adherence to High-Intensity Statins Following a Myocardial Infarction Hospitalization Among Medicare Beneficiaries. JAMA Cardiology. 2017;2(8):890–895.

30. Rosenson RS, Farkouh ME, Mefford M, et al. Trends in Use of High-Intensity Statin Therapy After Myocardial Infarction, 2011 to 2014. J Am Coll Cardiol. 2017;69(22):2696–2706.

31. Booth JN3, Colantonio LD, Chen L, et al. Statin Discontinuation, Reinitiation, and Persistence Patterns Among Medicare Beneficiaries After Myocardial Infarction: A Cohort Study. Circulation.Cardiovascular Quality & Outcomes. 2017;10(10):Ot.

32. Hickson RP, Robinson JG, Annis IE, Killeya-Jones LA, Fang G. It’s Not Too Late to Improve Statin Adherence: Association Between Changes in Statin Adherence from Before to After Acute Myocardial Infarction and All-Cause Mortality. Journal of the American Heart Association. 2019;8(7):e011378.

33. Cannon CP, de Lemos JA, Rosenson RS, et al. Use of Lipid-Lowering Therapies Over 2 Years in GOULD, a Registry of Patients With Atherosclerotic Cardiovascular Disease in the US. JAMA Cardiology. 2021.

34. Escudero-Sanchez G, Rico-Martin S, Sanchez-Bacaicoa C, et al. Optimal Control of all Modifiable Vascular Risk Factors Among Patients With Atherosclerotic Disease. A Real-Life Study. Curr Probl Cardiol. 2022;48(3):101530.

35. Boden WE, O’Rourke RA, Teo KK, et al. Optimal Medical Therapy with or without PCI for Stable Coronary Disease. The New England Journal of Medicine. 2007;356(15):1503– 1516. doi:10.1056/NEJMoa070829

36. Mendis S, Abegunde D, Yusuf S, et al. WHO study on Prevention of REcurrences of Myocardial Infarction and StrokE (WHO-PREMISE). Bull World Health Organ. 2005;83(11):820–829

37. Frye, August P, Brooks MM, et al. A Randomized Trial of Therapies for Type 2 Diabetes and Coronary Artery Disease The BARI 2D Study Group* A bs tr ac t. doi:10.1056/NEJMoa0805796)

38. Toth PP, Granowitz C, Hull M, Liassou D, Anderson A, Philip S. High Triglycerides Are Associated With Increased Cardiovascular Events, Medical Costs, and Resource Use: A Real-World Administrative Claims Analysis of Statin-Treated Patients With High Residual Cardiovascular Risk. Journal of the American Heart Association. 2018;7(15):e008740

39. Nichols GA, Philip S, Reynolds K, Granowitz CB, Fazio S. Increased residual cardiovascular risk in patients with diabetes and high versus normal triglycerides despite statin-controlled LDL cholesterol. Diabetes Obes Metab. 2019;21(2):366–371

40. Nichols GA, Philip S, Reynolds K, Granowitz CB, Fazio S. Increased Cardiovascular Risk in Hypertriglyceridemic Patients With Statin-Controlled LDL Cholesterol. Journal of Clinical Endocrinology & Metabolism. 2018;103(8):3019–3027

41. Klempfner R, Erez A, Sagit B, et al. Elevated Triglyceride Level Is Independently Associated With Increased All-Cause Mortality in Patients With Established Coronary Heart Disease: Twenty-Two-Year Follow-Up of the Bezafibrate Infarction Prevention Study and Registry. Circulation.Cardiovascular Quality & Outcomes. 2016;9(2):100–108

42. Libby P. Triglycerides on the rise: should we swap seats on the seesaw? Eur Heart J. 2015;36(13):774–776

43. Bhatt DL, Steg PG, Miller M, et al. Cardiovascular Risk Reduction with Icosapent Ethyl for Hypertriglyceridemia. N Engl J Med. 2019;380(1):11–22

44. Virani SS, Newby LK, Arnold SV, et al. 2023 AHA/ACC/ACCP/ASPC/NLA/PCNA Guideline for the Management of Patients With Chronic Coronary Disease: A Report of the American Heart Association/American College of Cardiology Joint Committee on Clinical Practice Guidelines. J Am Coll Cardiol. 2023;82(9):833–955. doi:10.1016/j.jacc.2023.04.003

45. Mondesir FL, Brown TM, Muntner P, et al. Diabetes, diabetes severity, and coronary heart disease risk equivalence: REasons for Geographic and Racial Differences in Stroke (REGARDS). Am Heart J. 2016;181:43–51.

46. Emerging Risk Factors Collaboration, Sarwar N, Gao P, et al. Diabetes mellitus, fasting blood glucose concentration, and risk of vascular disease: a collaborative meta-analysis of 102 prospective studies. Lancet. 2010;375(9733):2215–2222.

47. Cosentino F, Grant PJ, Aboyans V, et al. 2019 ESC Guidelines on diabetes, pre-diabetes, and cardiovascular diseases developed in collaboration with the EASD. Eur Heart J. 2020;41(2):255–323. doi:10.1093/eurheartj/ehz486

48. Marx N, Husain M, Lehrke M, Verma S, Sattar N. GLP-1 Receptor Agonists for the Reduction of Atherosclerotic Cardiovascular Risk in Patients With Type 2 Diabetes. Circulation. 2022;146(24):1882–1894. doi:10.1161/CIRCULATIONAHA.122.059595

49. Marso SP, Daniels GH, Brown-Frandsen K, et al. Liraglutide and Cardiovascular Outcomes in Type 2 Diabetes. N Engl J Med. 2016;375(4):311–322. doi:10.1056/NEJMoa1603827

50. Lincoff AM, Brown-Frandsen K, Colhoun HM, et al. Semaglutide and Cardiovascular Outcomes in Obesity without Diabetes. N Engl J Med. 2023;389(24):2221–2232. doi:10.1056/NEJMoa2307563

